# Osteoprotegerin (OPG) and its ligands RANKL and TRAIL in falciparum, vivax and knowlesi malaria: correlations with disease severity, and B cell production of OPG

**DOI:** 10.1101/2024.07.22.24310838

**Authors:** Arya Sheela Nair, John Woodford, Jessica Loughland, Dean Andrew, Kim Piera, Fiona Amante, Timothy William, Matthew J Grigg, James S McCarthy, Nicholas M Anstey, Michelle J Boyle, Bridget E Barber

**Affiliations:** QIMR Berghofer Medical Research Institute, Brisbane, Australia; Laboratory of Malaria Immunology and Vaccinology, National Institute of Allergy and Infectious Diseases, Bethesda, United States; Menzies School of Health Research, Darwin, Australia; Subang Jaya Medical Centre, Selangor, Malaysia; Peter Doherty Institute for Infection and Immunity, Melbourne, Australia; Burnet Institute, Melbourne, Australia

**Keywords:** Malaria, falciparum, vivax, knowlesi, osteoprotegerin, OPG, receptor activator of NF-ƙB ligand, RANKL, TNF-related apoptosis-inducing ligand, TRAIL

## Abstract

Osteoprotegerin (OPG) is a soluble decoy receptor for receptor activator of NF-ƙB ligand (RANKL) and TNF-related apoptosis-inducing ligand (TRAIL), and is increasingly recognised as a marker of poor prognosis in a number of diseases. Here we demonstrate that in Malaysian adults with falciparum and vivax malaria, OPG is increased, and its ligands TRAIL and RANKL decreased, in proportion to disease severity. In volunteers experimentally infected with *P. falciparum* and *P. vivax*, RANKL was suppressed, while TRAIL was unexpectedly increased, suggesting binding of OPG to RANKL prior to TRAIL. We also demonstrate that *P. falciparum* stimulates B cells to produce OPG *in vitro*, and that B cell OPG production is increased *ex vivo* in patients with falciparum, vivax and knowlesi malaria. Our findings provide further evidence of the importance of the OPG/RANKL/TRAIL pathway in pathogenesis of diseases involving systemic inflammation, and may have implications for adjunctive therapies. Further evaluation of the role of B cell production of OPG in host responses to malaria and other inflammatory diseases is warranted.

## Introduction

Osteoprotegerin (OPG) is a naturally circulating soluble decoy receptor for receptor activator of NF- ƙB ligand (RANKL) and TNF-related apoptosis-inducing ligand (TRAIL). OPG was initially identified as a key modulator of bone metabolism, produced by osteoblasts and regulating bone remodelling through its inhibition of RANKL-mediated osteoclastogenesis [1,2]. OPG however is also produced by vascular smooth muscle cells [3], endothelial cells [4], and immune cells including B lymphocytes and dendritic cells [5], and its role in the cardiovascular and immune systems is increasingly recognised [6]. OPG is now well documented to be a marker of poor prognosis in cardiovascular diseases, and has been associated with all-cause mortality in a number of studies [7,8]. The pathogenic effects of OPG within the cardiovascular system are incompletely understood, but are thought to relate in part to endothelial activation and dysfunction, mediated through upregulation of endothelial cell adhesion molecules, increased production of reactive oxygen species, apoptosis of human endothelial progenitor cells, and inhibition of endothelial nitric oxide (NO) synthase [7].

Many of the pathogenic processes mediated by OPG are also key features of severe malaria [9]. Indeed, OPG has recently been shown to be elevated in African children with *Plasmodium falciparum* malaria [10], in volunteers experimentally infected with *P. falciparum* and *P. vivax* [11], and in adults with malaria from the zoonotic parasite *P. knowlesi* [12]. In the latter, OPG was higher in those with severe disease, and was associated with adverse effects such as microvascular dysfunction and acute kidney injury, supporting a role in disease pathogenesis [12]. However, OPG has not been evaluated in adults with clinical malaria from *P. falciparum* and *P. vivax*, the most common causes of malaria worldwide. Furthermore, the cellular sources of OPG in malaria have not been investigated.

To extend our previous studies of OPG in knowlesi malaria [12], we evaluated plasma concentrations of OPG in Malaysian adults with severe and non-severe falciparum and vivax malaria. We also evaluated plasma concentrations of the OPG ligands RANKL and TRAIL in patients with falciparum, vivax and knowlesi malaria, and in volunteers experimentally infected with *P. falciparum* and *P. vivax*. Given previous studies demonstrating B cell production of OPG [5], we evaluated B cell OPG production following parasite stimulation *in vitro*, and measured B cell OPG production *ex vivo* in patients with falciparum, vivax and knowlesi malaria. Our data demonstrate that OPG is increased in proportion to disease severity, and RANKL and TRAIL decreased, in clinical malaria from all species, adding to the body of evidence supporting the importance of the OPG/RANKL/TRAIL pathway in pathogenesis of a range of diseases. We also demonstrate B cell production of OPG in malaria, highlighting the importance of the interaction between *Plasmodium* parasites and B cells, and suggesting that OPG may play a role in immune responses to malaria.

## Results

### OPG is increased in Malaysian patients with falciparum and vivax malaria

OPG was evaluated in 427 patients with malaria and 50 healthy controls enrolled at the same study site. Patients with malaria included 169 with *P. falciparum* (150 non-severe, 19 severe), 61 with *P. vivax* (53 non-severe, 8 severe), and 197 with *P. knowlesi* (149 non-severe, 48 severe). Clinical and pathophysiological data from this cohort of patients have been previously reported [13–15]. Baseline demographic and clinical characteristics are shown in **Table 1**. Overall, 328 (76%) of the patients with malaria were male, and the median age was 34 years (range 13 – 94 years).

**Table 1.** Baseline characteristics of Malaysian malaria patients and controls.

|  | **Controls (n=50)** | ***P. falciparum* (n=169)** | ***P. vivax* (n=61)** | ***P. knowlesi* (n=197)** |
| --- | --- | --- | --- | --- |
| Age, years (median, range) | 43 (14 – 69) | 27 (13 – 78) | 27 (13 – 79) | 45 (13-94) |
| Male sex, n (%) | 34 (68) | 124 (73) | 47 (77) | 153 (78) |
| Fever duration, days |  | 5 (3 – 7) | 5 (3 – 7) | 5 (3 – 7) |
| Parasite count, parasites/μL | | 10,500 (3425 – 33,571) | 4693 (2848 – 10,336) | 5550 (1596 – 32,455) |
| Severe malaria, n (%) |  | 19 (11%) | 8 (13%) | 48 (24%) |
| Creatinine, μmol/L | | 87 (73 – 103) | 87 (69 – 104) | 101 (81 – 122) |
| Lactate, mmol/L |  | 1.24 (0.91 – 1.66)<br>N=144 | 1.21 (0.83 – 1.56)<br>N=56 | 1.2 (0.97 – 1.61)<br>N=178 |
| Angiopoietin-2, pg/ml | 1182 (875 – 1597) | 3700 (2228- 5840) | 4986 (3580 – 6697) | 5083 (3297 – 8127) |
| VWF, pg/ml | 1156 (843 – 1634)<br>N=30 | 5470 (3906 – 7424)<br>N=54 | 4470 (3108 – 5561)<br>N=37 | 5171 (4251 – 6256)<br>N=85 |
| P-selectin, pg/ml | 40 (31 – 52) | 46 (35 – 69) | 47 (34 – 62) | 33 (25 – 42)<br>N=195 |
| ICAM-1, pg/ml | 149 (123 – 167) | 550 (395 – 705) | 507 (416 – 686) | 480 (377 – 644) |
| E-selectin, pg/ml | 19 (13 – 25) | 59 (42 – 86) | 69 (43 – 97) | 52 (40 – 68) |
| OPG, pg/ml | 986 (625 – 1463) | 2344 (1310 – 4666) | 2578 (1301 – 5642) | 2483 (1729 – 4265)<br>N=195 |
| RANKL<sup>a</sup>, pmol/L | 0.225 (0.141 – 0.335)<br>N=20 | 0.031 (0.031 – 0.083)<br>N=43 | 0.031 (0.031 – 0.031)<br>N=25 | 0.031 (0.031 – 0.031)<br>N=28 |
| TRAIL, pg/ml | 74 (66 – 99)<br>N=20 | 58 (45 – 84) | 58 (45 – 72) | 50 (36 – 63) |

Numbers are median (inter-quartile range) unless otherwise stated. HRP2: histidine rich protein 2; VWF: von Willibrand factor; ICAM-1: intracerebral adhesion molecule 1; OPG: osteoprotegerin; RANKL: receptor activator of NF- $\kappa$ B ligand; TRAIL: tumour necrosis factor apoptosis inducing ligand.
<sup>a</sup>Patients with a RANKL concentration below the limit of detection were assigned a value half that of the limit of detection, which was 0.0625 pg/ml. RANKL was below the limit of detection in 27 (63%) patients with *P. falciparum*, 21 (84%) patients with *P. vivax* and 22 (79%) patients with *P. knowlesi*.

OPG was increased in patients with falciparum and vivax malaria, compared to controls (p<0.0001 for both comparisons; **Table 1** and **Figure 1**). OPG was also higher in patients with severe compared to non-severe disease, although in vivax malaria this difference was not statistically significant (p=0.052; **Figure 1**). There was no significant difference in the concentration of plasma OPG between species. There was also no significant difference in concentration of OPG between males and females, in either falciparum or vivax malaria.

**Figure 1.**
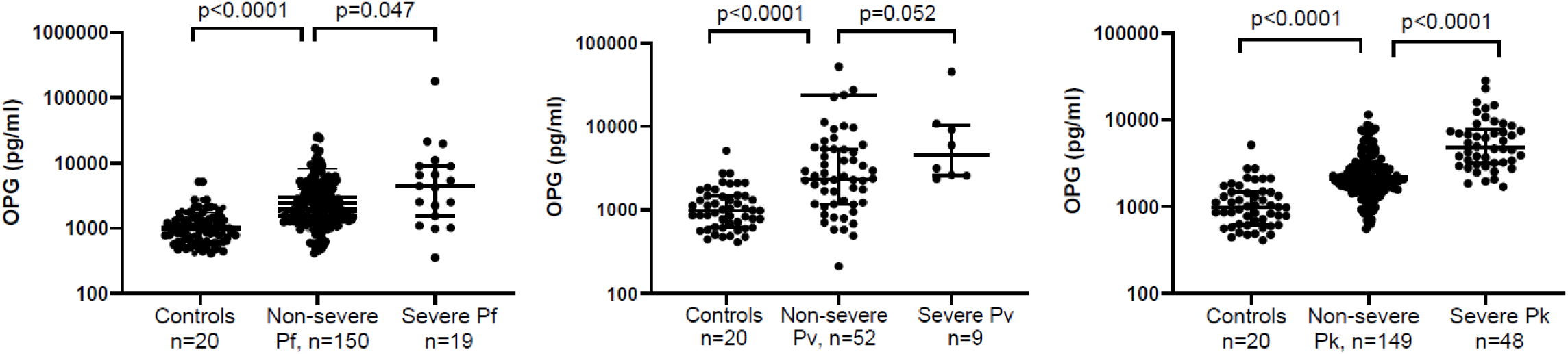
Plasma concentrations of osteoprotegerin (OPG) in Malaysian patients with *P. falciparum*, *P. vivax* and *P. knowlesi* malaria. Pf = *P. falciparum*; Pv = *P. vivax*; Pk = *P. knowlesi*. Errors bars represent median and inter-quartile range. Data for patients with knowlesi malaria have been previously reported [12], and are included here for comparison with falciparum and vivax malaria.

In patients with falciparum malaria, OPG was correlated with parasitaemia and age (**Table 2**). In falciparum malaria OPG also correlated with other markers of disease severity including creatinine and lactate, and endothelial activation markers angiopoietin-2, intercellular adhesion molecule 1 (ICAM-1) and E-selectin, after controlling for age and parasitemia. Interestingly, in falciparum malaria there was no correlation between OPG and the parasite biomass marker HRP2 (r=0.06, p=0.47), despite a correlation with HRP2 being observed for the other endothelial Weibel-Palade body (WPB) constituents, angiopoietin-2 (r=0.41, p<0.0001) and VWF (r=0.29, p=0.032).

**Table 2.** Correlations with OPG in patients with falciparum and vivax malaria.

|  | *P. falciparum* (n=169) |  |  |  | *P. vivax* (n=61) |  |  |  |
| --- | --- | --- | --- | --- | --- | --- | --- | --- |
|  | Univariate analysis |  | Controlling for parasitemia and age |  | Univariate analysis |  | Controlling for parasitemia and age |  |
|  | Correlation coefficient | P value | Correlation coefficient | P value | Correlation coefficient | P value | Correlation coefficient | P value |
| Parasite count | 0.29 | <0.0001 | 0.29<sup>a</sup> | 0.001 | 0.47 | 0.0002 | 0.42<sup>a</sup> | 0.0008 |
| Age | 0.24 | 0.002 | 0.20<sup>b</sup> | 0.008 | 0.05 | 0.67 |  |  |
| HRP2<sup>c</sup> | 0.06 | 0.47 |  |  |  |  |  |  |
| Creatinine | 0.24 | 0.002 | 0.18 | 0.020 | 0.05 | 0.68 |  |  |
| Lactate | 0.18 | 0.031 | 0.25 | 0.002 | 0.30 | 0.026 | 0.28 | 0.038 |
| IL-6 | 0.47 | <0.0001 | 0.39 | <0.0001 | 0.61 | <0.0001 | 0.47 | 0.0002 |
| Angiopoietin-2 | 0.22 | 0.004 | 0.17 | 0.022 | 0.40 | 0.001 | 0.24 | 0.07 |
| vWF<sup>d</sup> | 0.30 | 0.026 | 0.14 | 0.33 | 0.40 | 0.014 | 0.29 | 0.10 |
| P-selectin | 0.05 | 0.54 |  |  | 0.09 | 0.51 |  |  |
| ICAM-1 | 0.17 | 0.023 | 0.23 | 0.003 | 0.19 | 0.15 |  |  |
| E-selectin | 0.41 | <0.0001 | 0.37 | <0.0001 | 0.39 | 0.002 | 0.38 | 0.003 |
| Hb nadir | 0.12 | 0.11 |  |  | -0.01 | 0.91 |  |  |
| Plt nadir | -0.32 | <0.0001 | -0.29 | 0.0001 | -0.20 | 0.12 |  |  |
| Endothelial function<sup>e</sup> | 0.01 | 0.96 |  |  | -0.01 | 0.95 |  |  |

<sup>a</sup>controlling for age; <sup>b</sup>controlling for parasitemia; <sup>c</sup>measured in 153 patients; <sup>d</sup>measured in 144 patients with *P. falciparum*, 56 with *P. vivax*, and 178 patients with *P. knowlesi* malaria; <sup>e</sup>measured by peripheral arterial tonometry in 88 patients with *P. falciparum* and 38 patients with *P. vivax*. HRP2: histidine rich protein 2; vWF: von Willebrand Factor; ICAM-1: intracerebral adhesion molecule 1; Hb: haemoglobin; Plt: platelet

In patients with vivax malaria, OPG was correlated with parasitaemia, lactate, angiopoietin-2, vWF and E-selectin, on univariate analysis (**Table 2**). The correlations with lactate and E-selectin remained significant after controlling for age and parasitemia. There was no correlation between OPG and endothelial function, in either falciparum or vivax malaria.

### RANKL and TRAIL are reduced in Malaysian patients with falciparum, vivax and knowlesi malaria

Plasma concentrations of the OPG ligand TRAIL were measured in all patients with falciparum, vivax and knowlesi malaria, while RANKL was measured in a subset of patients including 43 with *P. falciparum* (24 non-severe, 19 severe), 25 with *P. vivax* (19 non-severe, 6 severe), and 28 with *P. knowlesi* (15 non-severe, 13 severe). Consistent with the binding of OPG to its ligands, plasma concentrations of RANKL and TRAIL were both reduced in patients with malaria compared to controls (**Table 1** and **Figure 2**). RANKL was suppressed to below the level of detection in 70/96 (73%) of all malaria patients, including 27 (63%) of those with *P. falciparum*, 21 (84%) of those with *P. vivax*, and 22 (79%) of those with *P. knowlesi*, compared to only 1 of 20 (5%) controls (p<0.0001 for all comparisons between each species and controls). RANKL was undetectable in all patients with severe knowlesi and vivax malaria, although was detectable (at mostly low levels) in 8/19 (42%) of patients with severe falciparum malaria. Similar to RANKL, TRAIL was also reduced in patients with falciparum, vivax and knowlesi malaria compared to controls (p=0.007, p=0.0004, and p<0.0001, respectively). In patients with vivax and knowlesi malaria, TRAIL was lower in those with severe compared to non-severe disease (p=0.0009 and p<0.0001, respectively); however, this difference was not observed in patients with falciparum malaria. There were no differences in concentrations of RANKL or TRAIL between males and females, in any species.

**Figure 2.**
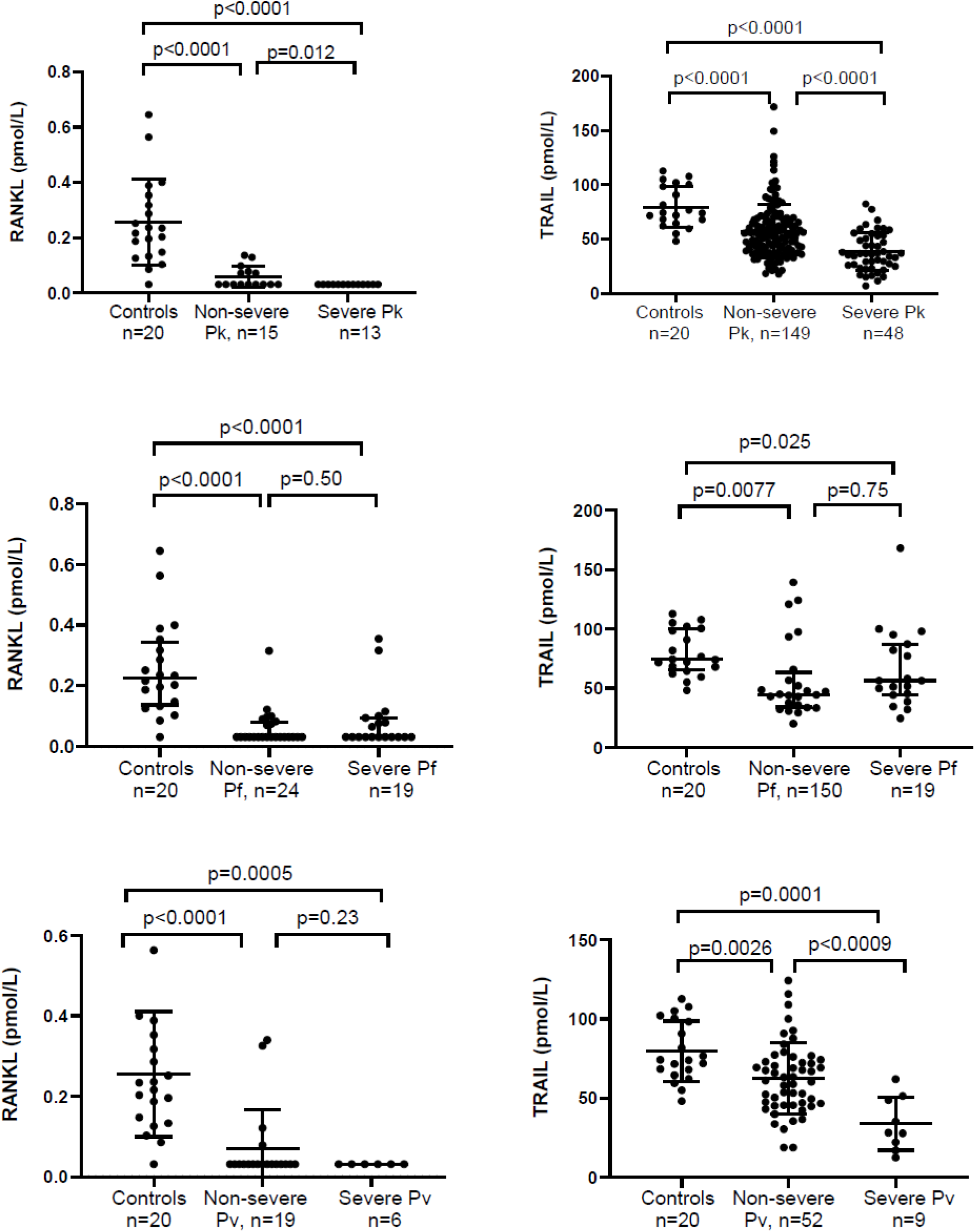
Plasma concentrations of RANKL and TRAIL in Malaysian patients with *P. falciparum*, *P. vivax* and *P. knowlesi* malaria. Errors bars represent median and inter- quartile range.

In all species, TRAIL was inversely correlated with parasitaemia and with age, although in *P. falciparum* and *P. knowlesi* the correlation with age did not remain significant after controlling for parasitaemia (**Table 3**). As expected (due to the binding of TRAIL to OPG), in all species TRAIL was inversely correlated with OPG; in *P. vivax* and *P. knowlesi* this remained significant after controlling for age and parasitaemia (**Table 3**). In all species TRAIL was also inversely correlated with angiopoietin-2, after controlling for age and parasitaemia, and in *P. knowlesi* this association remained significant after controlling for OPG. In *P. knowlesi* TRAIL was inversely correlated with creatinine, and positively correlated with haemoglobin and platelet nadir, with these correlations remaining significant after controlling for age, parasitaemia and OPG. In patients with *P. knowlesi*, in a backward stepwise linear regression model including age, parasitaemia, and OPG, TRAIL was inversely proportional to risk of having acute kidney injury on admission (defined by KDIGO criteria; **Supplementary Table 1**). However, this did not remain significant if angiopoietin-2, another known risk factor for AKI in knowlesi malaria [12], was included in the model.

**Table 3.**
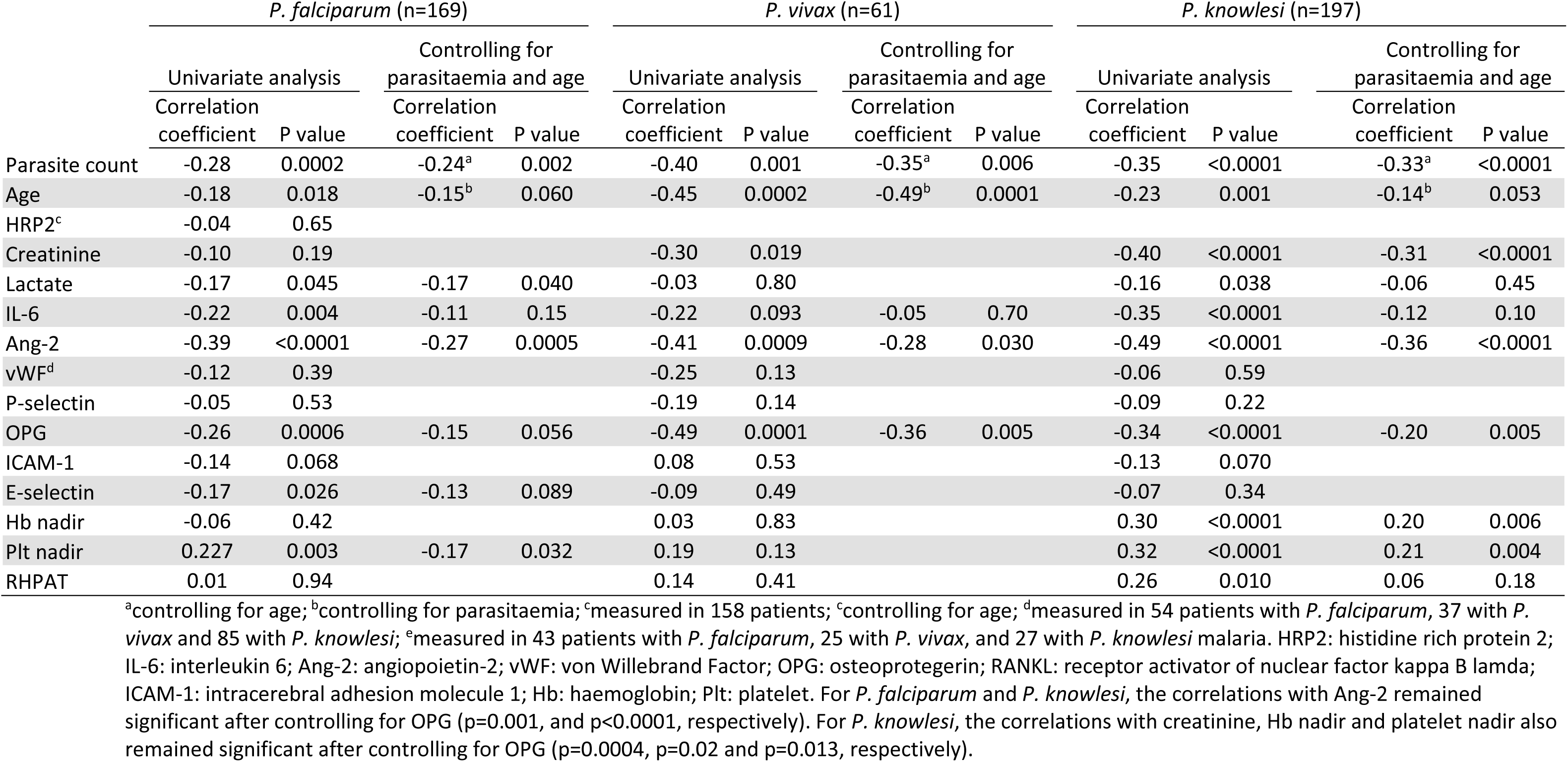
Correlations with TRAIL in patients with falciparum, vivax and knowlesi malaria.

|  | *P. falciparum* (n=169) |  |  |  | *P. vivax* (n=61) |  |  |  | *P. knowlesi* (n=197) |  |  |  |
| --- | --- | --- | --- | --- | --- | --- | --- | --- | --- | --- | --- | --- |
|  | Univariate analysis |  | Controlling for parasitaemia and age |  | Univariate analysis |  | Controlling for parasitaemia and age |  | Univariate analysis |  | Controlling for parasitaemia and age |  |
|  | Correlation coefficient | P value | Correlation coefficient | P value | Correlation coefficient | P value | Correlation coefficient | P value | Correlation coefficient | P value | Correlation coefficient | P value |
| Parasite count | -0.28 | 0.0002 | -0.24<sup>a</sup> | 0.002 | -0.40 | 0.001 | -0.35<sup>a</sup> | 0.006 | -0.35 | <0.0001 | -0.33<sup>a</sup> | <0.0001 |
| Age | -0.18 | 0.018 | -0.15<sup>b</sup> | 0.060 | -0.45 | 0.0002 | -0.49<sup>b</sup> | 0.0001 | -0.23 | 0.001 | -0.14<sup>b</sup> | 0.053 |
| HRP2<sup>c</sup> | -0.04 | 0.65 |  |  |  |  |  |  |  |  |  |  |
| Creatinine | -0.10 | 0.19 |  |  | -0.30 | 0.019 |  |  | -0.40 | <0.0001 | -0.31 | <0.0001 |
| Lactate | -0.17 | 0.045 | -0.17 | 0.040 | -0.03 | 0.80 |  |  | -0.16 | 0.038 | -0.06 | 0.45 |
| IL-6 | -0.22 | 0.004 | -0.11 | 0.15 | -0.22 | 0.093 | -0.05 | 0.70 | -0.35 | <0.0001 | -0.12 | 0.10 |
| Ang-2 | -0.39 | <0.0001 | -0.27 | 0.0005 | -0.41 | 0.0009 | -0.28 | 0.030 | -0.49 | <0.0001 | -0.36 | <0.0001 |
| vWF<sup>d</sup> | -0.12 | 0.39 |  |  | -0.25 | 0.13 |  |  | -0.06 | 0.59 |  |  |
| P-selectin | -0.05 | 0.53 |  |  | -0.19 | 0.14 |  |  | -0.09 | 0.22 |  |  |
| OPG | -0.26 | 0.0006 | -0.15 | 0.056 | -0.49 | 0.0001 | -0.36 | 0.005 | -0.34 | <0.0001 | -0.20 | 0.005 |
| ICAM-1 | -0.14 | 0.068 |  |  | 0.08 | 0.53 |  |  | -0.13 | 0.070 |  |  |
| E-selectin | -0.17 | 0.026 | -0.13 | 0.089 | -0.09 | 0.49 |  |  | -0.07 | 0.34 |  |  |
| Hb nadir | -0.06 | 0.42 |  |  | 0.03 | 0.83 |  |  | 0.30 | <0.0001 | 0.20 | 0.006 |
| Plt nadir | 0.227 | 0.003 | -0.17 | 0.032 | 0.19 | 0.13 |  |  | 0.32 | <0.0001 | 0.21 | 0.004 |
| RHPAT | 0.01 | 0.94 |  |  | 0.14 | 0.41 |  |  | 0.26 | 0.010 | 0.06 | 0.18 |

<sup>a</sup>controlling for age; <sup>b</sup>controlling for parasitaemia; <sup>c</sup>measured in 158 patients; <sup>c</sup>controlling for age; <sup>d</sup>measured in 54 patients with *P. falciparum*, 37 with *P. vivax* and 85 with *P. knowlesi*; <sup>e</sup>measured in 43 patients with *P. falciparum*, 25 with *P. vivax*, and 27 with *P. knowlesi* malaria. HRP2: histidine rich protein 2; IL-6: interleukin 6; Ang-2: angiopoietin-2; vWF: von Willebrand Factor; OPG: osteoprotegerin; RANKL: receptor activator of nuclear factor kappa B lamda; ICAM-1: intracerebral adhesion molecule 1; Hb: haemoglobin; Plt: platelet. For *P. falciparum* and *P. knowlesi*, the correlations with Ang-2 remained significant after controlling for OPG (p=0.001, and p<0.0001, respectively). For *P. knowlesi*, the correlations with creatinine, Hb nadir and platelet nadir also remained significant after controlling for OPG (p=0.0004, p=0.02 and p=0.013, respectively).

Previous studies have demonstrated that the OPG/TRAIL ratio may have greater prognostic value than either biomarker alone [7], and we therefore also evaluated correlates of this ratio. In general, associations with other biomarkers of severity were indeed stronger with the OPG/TRAIL ratio than with either TRAIL or OPG alone (**Supplementary Table 2)**. In each malaria species, the OPG/TRAIL ratio was associated with IL-6, angiopoietin-2 and E-selectin, after controlling for age and parasitemia. In falciparum and knowlesi malaria, the OPG/TRAIL ratio was also correlated with lactate, ICAM-1 and platelet nadir, after controlling for age and parasitemia.

Clinical correlates of RANKL were not evaluated due to the low proportion of patients with detectable RANKL. However, given previous data from murine studies demonstrating increased RANKL expression following recovery from acute malaria (resulting in increased osteoclastogenesis and bone resorption) [16], we measured RANKL on samples taken on day 28 follow-up in 15 patients with *P. falciparum*, 15 with *P. vivax*, and 17 with *P. knowlesi* malaria. In all species, RANKL concentrations at day 28 were increased above enrolment levels (p<0.001 for each species), but were not elevated above the levels observed in healthy controls (**Supplementary Figure 1**).

### RANKL is reduced, while TRAIL increased, in volunteers infected with *P. falciparum* and *P. vivax*

Characteristics of participants enrolled in malaria volunteer infection studies (VIS), and details of these studies, are described in **Supplementary Table 3.** Data were included from 28 participants enrolled in *P. falciparum* VIS, and 16 participants enrolled in *P. vivax* VIS.

We previously reported that concentrations of OPG were increased early in volunteers experimentally infected with *P. falciparum* and *P. vivax*, consistent with the increase observed in Malaysian patients with clinical malaria [11]. Also consistent with data from the Malaysian patients with malaria, RANKL was reduced in volunteers infected with *P. falciparum* and *P. vivax*, being suppressed to below the level of detection in 7/28 (25%) participants infected with *P. falciparum*, and 7/16 (44%) participants infected with *P. vivax* **(Figure 3)**. In both the *P. falciparum* and the *P. vivax* groups, RANKL was lower at the time of antimalarial treatment compared to baseline (p<0.0001 and p=0.0013 respectively). By the end of the studies, RANKL had returned to baseline levels in the *P. vivax* group, although remained slightly lower than baseline levels in the *P. falciparum* group (p=0.042). On the day of treatment, RANKL was inversely correlated with parasitemia in participants inoculated with *P. vivax*, although this did not remain significant after controlling for OPG (**Supplementary Table 4**). There was no correlation between RANKL and parasitemia in participants inoculated with *P. falciparum*. As expected, on the day of treatment RANKL was strongly inversely correlated with OPG in both the *P. falciparum* (r=-0.66, p=0.0002) and *P. viva*x (r=-0.92, p=<0.0001) groups, remaining significant after controlling for parasitemia.

**Figure 3.**
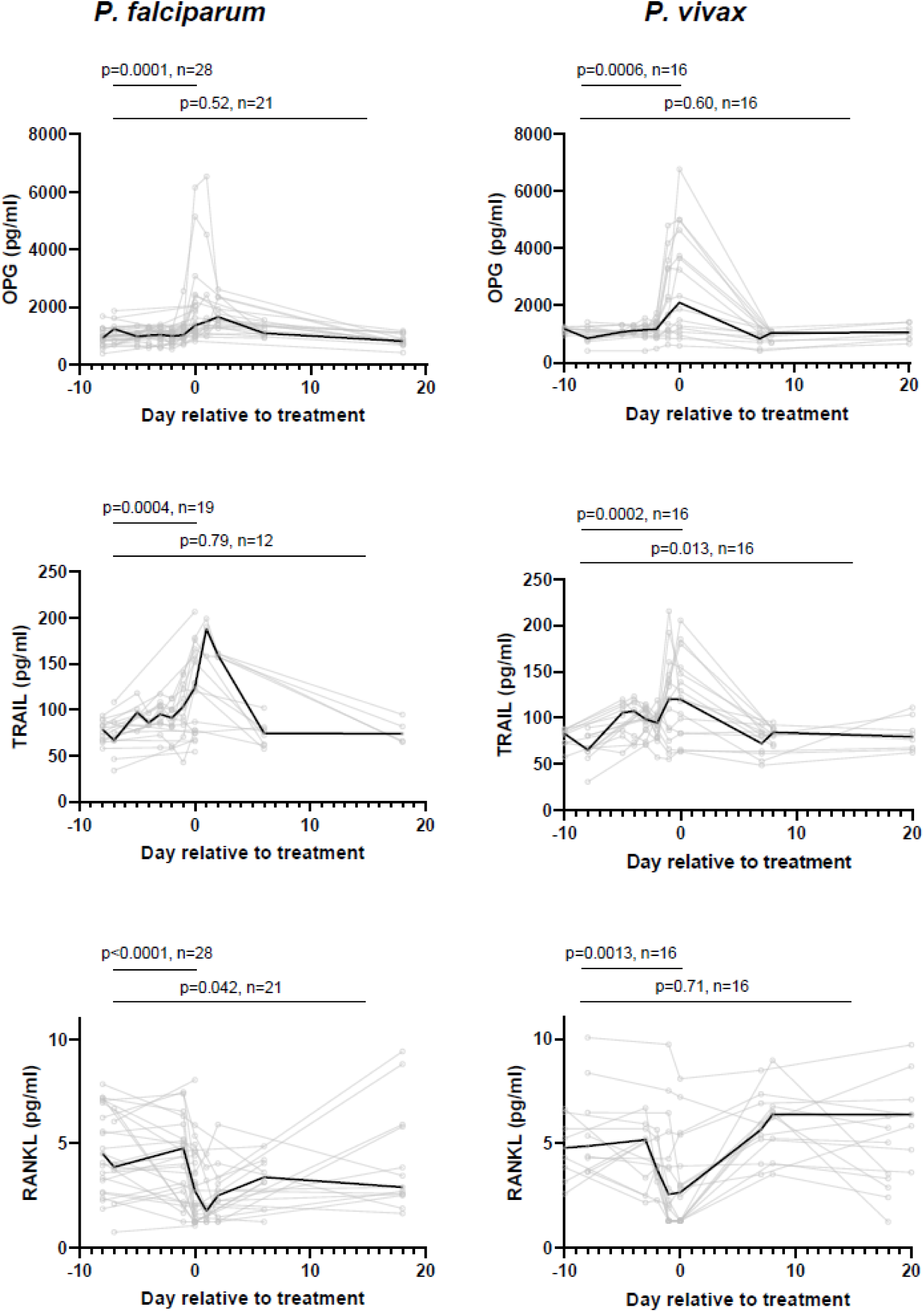
Longitudinal concentrations of OPG, RANKL and TRAIL participants inoculated with *P. falciparum* (left column) and *P. vivax* (right column). Black line represents median. Grey lines represent individual participants. Comparisons by Wilcoxon matched pair signed rank test for baseline (day of controlled malaria infection) and day of antimalarial treatment, and baseline and end of study measurements. Data for OPG has already been reported, and is included here for context.

In contrast to Malaysian patients with clinical malaria, plasma concentrations of TRAIL were unexpectedly increased in volunteers infected with *P. falciparum* and *P. vivax*, with concentrations peaking on the day of treatment (*P. falciparum:* p=0.0004, *P. vivax:* p=0.0002) before falling to near baseline levels following treatment (**Figure 3**). On the day of treatment TRAIL was positively correlated with parasitemia in participants inoculated with *P. vivax*, but not *P. falciparum* (**Supplementary Table 5**).

### B cell production of OPG is increased following stimulation by parasitised red blood cells

Given the marked elevation in OPG observed in experimental and clinical malaria, and the correlations with disease severity in the latter, we sought to determine if *P. falciparum* parasites could stimulate B cells to produce OPG. Peripheral blood mononuclear cells (PBMCs) isolated from buffy coats from healthy donors (n=10) from Australian Red Cross LifeBlood were stimulated with *P. falciparum* pRBCs or uRBCs. Expression of OPG from B cells was increased in donors after stimulation with pRBCs, compared to uRBCs (p=0.0039, **Figure 4A/B**). Negligible OPG was detected from other cell lineages, including NK cells and monocytes **(Supplementary Figure 2**). To identify the B cell subsets producing OPG in response to parasite stimulation, B cells were characterised as naïve (IgD+) or non-naïve (IgD-) B cells (**Figure 4C)**. Following stimulation with pRBCs, the large majority of OPG was produced by naïve (IgD+) B cells (**Figure 4D**).

**Figure 4.**
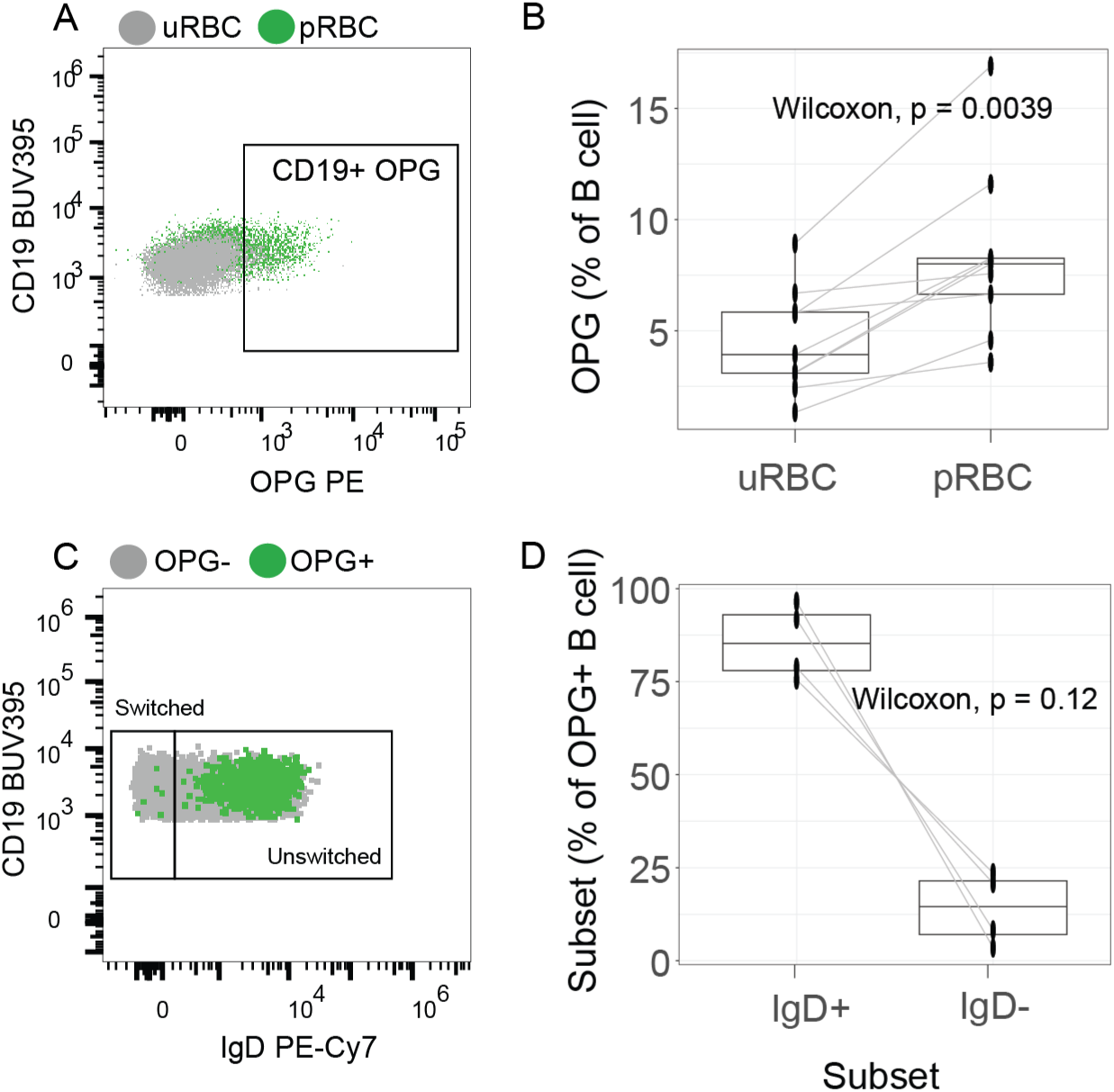
Malaria parasites can induce OPG expression in B cells. **A)** PBMCs from buffy from Australian donors (n=10) were stimulated with uninfected or parasite infected RBCs and OPG production from B cells assessed by flow cytometry. Representative gating figure of OPG from B cells following pRBC or uRBC stimulation. **B)** OPG B cell expression from malaria-naïve donors, after stimulation with uninfected RBCs (uRBCs) or *P. falciparum*-infection RBCs (pRBCs). P is Wilcoxon signed-rank test.**; C)** Analysis of B cell subsets that produce OPG. Representative gating figure of IgD expression from OPG positive cells B cells. **D)** IgD+ and IgD- B cell subsets as a proportion of all OPG+ B cells following parasite stimulation of PBMCs from malaria naïve donors (n=4). P is Wilcoxon signed-rank test. See also Supplementary Figure 3.

### B cell production of OPG is increased during clinical malaria

To evaluate whether B cell expression of OPG is increased in clinical malaria, we measured *ex vivo* OPG production from B cells from Malaysian patients with malaria (6 with *P. falciparum*, 8 with *P. vivax* and 7 with *P. knowles*i) on enrolment and at day 28, and from 5 healthy controls recruited from the same study site. Age and sex were similar between groups (**Supplementary Table 6).** Compared to controls, *ex vivo* OPG B cell expression was higher in patients with falciparum, vivax and knowlesi malaria, although numbers were small and the latter did not reach statistical significance (p=0.017, p=0.045 and p=0.073, respectively; **Figure 5A/B, Supplementary Figure 3A**). There was limited OPG detected from other cell lineages (**Supplementary Figure 3B).** B cell expression of OPG had decreased by day 28 in patients with falciparum, vivax and knowlesi malaria (p=0.062, p=0.078, and p=0.016, respectively; **Figure 5B**). In contrast to parasite stimulated PMBCs from blood donors, in patients with malaria OPG from B cells was detected in both IgD+ and IgD- subsets, with no difference in subset distribution between species (**Figure 5C/D)**. Within IgD- cells, the majority of OPG B cells were CD27 positive/CD21 negative memory B cells.

**Figure 5.**
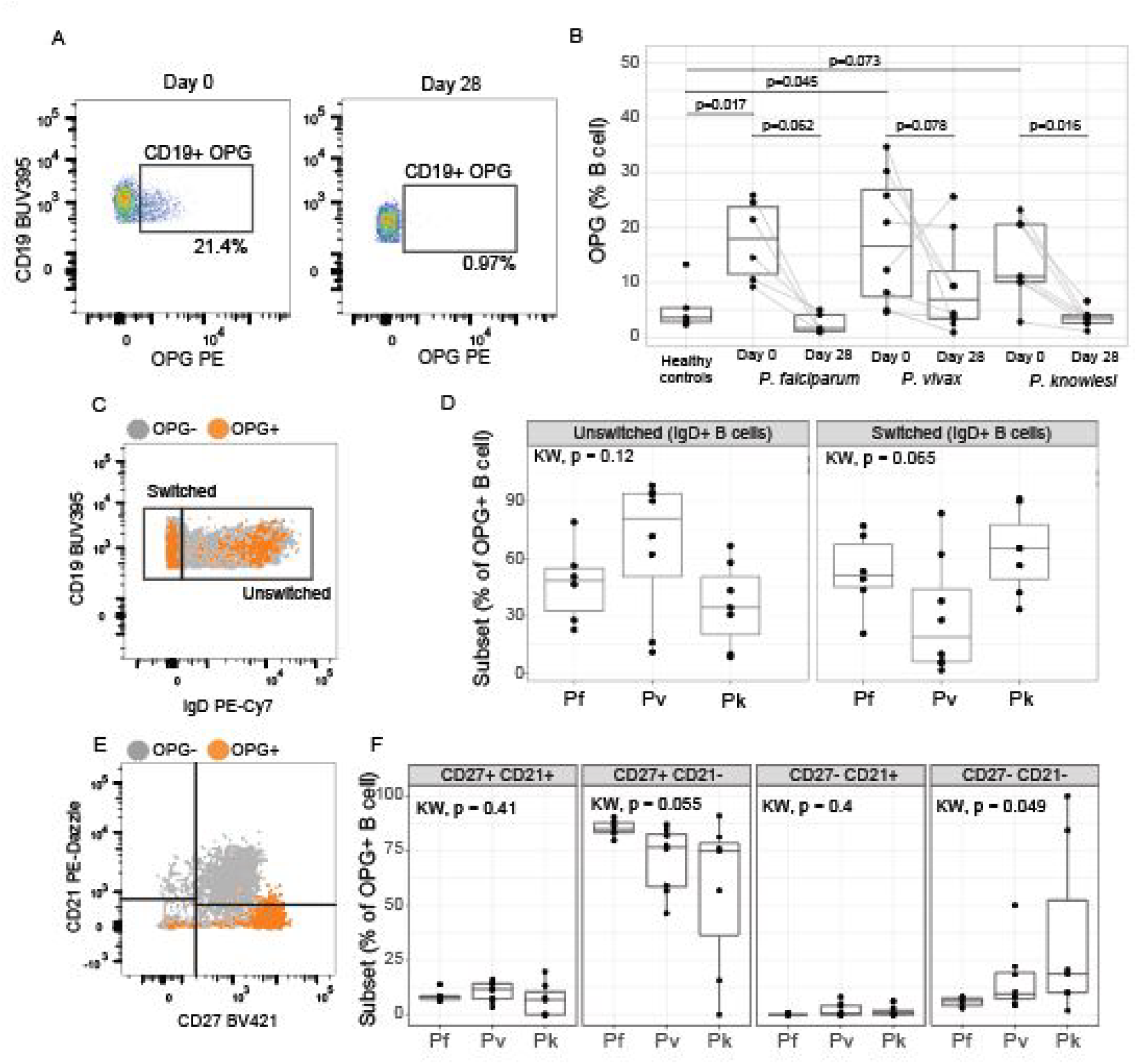
B cell production of OPG during clinical malaria. **A)** OPG expression from B cells at enrolment (day 0) and convalescence (day 28), gating example. **B)** Proportion of B cells expressing OPG in Malaysian patients with *P. falciparum* (n=6 day 0 and n=5 day 28), *P. vivax* (n=8) and *P. knowlesi* (n=7), compared to Malaysian healthy controls (n=5). P value between healthy controls and day 0 malaria infection calculated using Mann Whitney. P value between day 0 and day 28 calculated using Wilcox signed rank. **C)** IgD expression in OPG+ B cells, gating example. **D)** IgD+ and IgD- B cell subsets as a proportion of all OPG+ B cells at enrollment from malaria patients. P value was calculated using Kruskal-Wallis. **E)** CD27 and CD21 expression in OPG+ IgD- B cells, gating example. **F)** CD27 and CD21 B cell subsets as a proportion of all IgD- OPG+ B cells at enrollment from malaria patients. P value was calculated using Kruskal-Wallis. See also Supplementary Figure 3.

## Discussion

OPG is increasingly recognised as a marker of disease severity in a wide range of conditions. We previously reported that OPG was increased in adults with knowlesi malaria, and associated with markers of disease severity. In this study we extend these reports and demonstrate that OPG is also increased, and associated with disease severity, in Malaysian adults with falciparum and vivax malaria. We also demonstrate that the OPG ligands RANKL and TRAIL are reduced in adults with falciparum, vivax and knowlesi malaria, with this reduction in TRAIL being associated with markers of disease severity. Consistent with these findings in clinical malaria, RANKL was also reduced in volunteers enrolled in *P. falciparum* and *P. vivax* volunteer infection studies, although TRAIL was unexpectedly increased, suggesting that binding of OPG to RANKL precedes binding to TRAIL. Finally, we show that *P. falciparum* pRBCs stimulate B cell production of OPG *in vitro,* and that B cell production of OPG is increased *ex vivo* in patients with falciparum, vivax and knowlesi malaria.

Our finding that OPG is elevated in adults with falciparum malaria is consistent with a report of elevated OPG in African children with cerebral malaria [10]. In the current study, in adults with falciparum malaria OPG was associated with markers of inflammation and disease severity including IL-6, creatinine and lactate, and endothelial activation markers angiopoietin-2, E-selectin and ICAM-1, after controlling for age and parasitemia. These findings are consistent with the production of OPG from endothelial WPBs in response to inflammatory cytokines [4], and the role of OPG in inhibiting endothelial nitric oxide, exacerbating endothelial activation and upregulating adhesion molecules [7,17,18]. These findings suggest that OPG may play a key role in mediating pathogenesis of falciparum malaria, particularly given the independent role of endothelial activation in predicting severe disease and death in falciparum malaria [19].

OPG was also associated with markers of disease severity in patients with vivax malaria, including IL- 6, E-selectin, and lactate. This association between OPG and IL-6 in both falciparum and vivax malaria is consistent with murine data demonstrating that LPS-induced production of inflammatory cytokines was reduced in mice lacking OPG [20]. Pathogenic mechanisms in *P. vivax* are less well understood than in *P. falciparum* malaria [21]; however, the inflammatory response induced by *P. vivax* parasites has been shown to be more prominent than in falciparum malaria [22], and our data suggest that OPG may contribute to this inflammatory response.

To our knowledge this is the first study that has evaluated concentrations of the OPG ligands RANKL and TRAIL in patients with malaria from any species. Consistent with the binding of these ligands to OPG, concentrations of RANKL and TRAIL were both reduced in patients with clinical malaria. This was particularly apparent for RANKL, which was suppressed to below the level of detection in the majority of patients with malaria, even in those with non-severe disease. This suppression of RANKL would be expected to lead to reduced RANKL/RANK signaling, thereby increasing downstream production of proinflammatory cytokines [20].

As with RANKL, concentrations of TRAIL were also reduced in patients with falciparum, vivax, and knowlesi malaria. In vivax and knowlesi malaria, concentrations were lower in patients with severe compared to non-severe disease; however, this difference was not observed in patients with falciparum malaria. In all species, concentrations of TRAIL were inversely proportional to parasitemia, and with the endothelial activation marker angiopoietin-2. In knowlesi malaria, concentrations of TRAIL were associated with haemoglobin nadir, platelet nadir, and inversely with creatinine, after controlling for parasitemia and OPG, suggesting that reduced levels of TRAIL may contribute to disease pathogenesis. Low levels of TRAIL have also been associated with poor outcomes in cardiovascular disease [7]. While the mechanisms of the potential protective effects of TRAIL are uncertain, TRAIL has been shown to increase endothelial nitric oxide production, and to inhibit angiotensin II-induced production of reactive oxygen species, possibly leading to stabilisation of activated endothelium [23–25]. We found an association between TRAIL and endothelial function in patients with knowlesi malaria on univariate analysis; this association was also observed with the OPG/TRAIL ratio, with the latter remaining significant after controlling for parasitemia.

We also evaluated concentrations of RANKL and TRAIL in participants enrolled in *P. vivax* and *P. falciparum* malaria volunteer infection studies. Consistent with the suppression of RANKL in patients with clinical malaria, RANKL was also decreased on the day of treatment in these participants. However, in contrast to the decreased concentrations of TRAIL observed in patients with clinical malaria, TRAIL was unexpectedly increased in participants inoculated with the *P. falciparum* and *P. vivax*. This finding suggests that TRAIL increases early in infection, but declines as disease progresses, likely due to binding of TRAIL to increasing concentrations of OPG. The fact that suppression of RANKL precedes reductions in TRAIL is consistent with the greater binding affinity of OPG to RANKL [26], with OPG possibly only binding to TRAIL later in the disease course once free RANKL is no longer available.

Given the increase in OPG in clinical malaria and the likely role that this biomarker plays in a number of pathophysiological pathways, we sought to determine the role of immune cells in producing OPG in malaria. B cells are well-recognised as a source of OPG [5]; indeed, in a mouse study, Li et al. demonstrated that B cells accounted for >60% of all bone marrow production of OPG, and that B cell deficient mice had reduced levels of OPG [27]. In the current study, we found that *P. falciparum*- infected RBCs stimulated B cells to produce OPG *ex vivo*. Further, B cell expression of OPG was increased in patients with falciparum, vivax and knowlesi malaria, compared to healthy controls. While OPG producing B cells *in vitro* were largely naïve, during clinical infection both naïve and memory B cell subsets produced OPG, consistent with activation of the adaptive immune response. Our findings suggest that B cell production of OPG may be contributing at least in part to the substantial elevation in plasma OPG in patients with malaria. This is supported by the fact that in falciparum malaria, angiopoietin 2, the production of which is specific to endothelial cells, was associated with the parasite biomass marker HRP2, yet OPG was not. OPG was associated only with peripheral parasitemia, which may reflect admixture of circulating rather than sequestered parasites with B cells residing in lymph nodes, bone marrow, and the white pulp of the spleen, the latter perfused by the fast splenic circulation and with few sequestered parasites compared to the B cell-poor parasite-rich red pulp [28,29].

B cell production of OPG in malaria is also consistent with a growing body of evidence suggesting a link between OPG and immunity, with OPG shown to regulate B cell maturation and development of efficient antibody responses through CD40L signalling [5]. In a murine study, OPG deficient mice accumulated immature B cells in their spleens, and generated impaired antibody responses to a T-cell dependent antigen challenge [5]. Another study demonstrated that treatment of mice with OPG increased humoral immune responses to a pneumococcal vaccine [30]. It is therefore possible that in malaria, despite possible adverse pathogenic consequences of elevated OPG, B cell production of OPG may play a role in development of antimalarial immunity. Further studies evaluating associations between OPG and antimalarial immune responses are warranted.

Our study had limitations. First, the strength of many of the reported correlations with OPG and TRAIL was moderate, and it is possible that other confounding factors contributed to these associations. Second, although we controlled for parasitemia and age, we did not adjust for multiple correlations, and our findings should be interpreted accordingly. Third, for the experiments evaluating B cell production of OPG, sample sizes were small, limiting our ability to conduct statistical comparisons between groups. Finally, while we hypothesise that B cell production of OPG may be contributing to the elevated concentrations of plasma OPG in malaria, we did not evaluate other sources of OPG.

In summary, we found that plasma OPG was increased in adults with falciparum and vivax malaria, and associated with markers of disease severity in both species. The OPG ligands RANKL and TRAIL were both suppressed in clinical falciparum, vivax and knowlesi malaria, with reductions in TRAIL also associated with markers of severity, suggesting that suppression of RANKL and TRAIL may contribute to OPG-mediated pathogenesis in each *Plasmodium* species. In participants enrolled in malaria VIS, RANKL was also suppressed while TRAIL was unexpectedly increased, suggesting that OPG binds to RANKL early in disease and later to TRAIL as disease progresses. Finally, we demonstrate B cell production of OPG in clinical malaria, highlighting a potential role of OPG in the host immune response to malaria. Further studies evaluating the role of OPG in antimalarial immunity, and B cell production of OPG in other conditions, are warranted.

## Methods

### Malaysian study site, patients and procedures

Patients were enrolled as part of an observational study of all malaria patients admitted to Queen Elizabeth Hospital, an adult tertiary-referral hospital in Sabah, Malaysia [14]. For the current study, patients enrolled between September 2010 and December 2012 were included if they had PCR-confirmed *P. falciparum, P. vivax* or *P. knowlesi* monoinfection, were non-pregnant, ≥12 years old, had no major comorbidities or concurrent illness and were within 18 h of commencing antimalarial treatment. Parasite counts were determined by microscopy, and parasite species identified by PCR. Venous blood was collected in lithium heparin and citrate tubes for pathophysiological biomarkers on enrolment and at day 28. PBMCs were isolated and fractioned by Ficoll® Paque Plus (Cytiva) density-gradient centrifugation and frozen at -70° C in 10% dimethyl sulfoxide (DMSO) with 90% fetal bovine serum (FBS). Severe malaria was defined according to modified WHO criteria [31], as previously described [14]. Healthy controls were visitors or relatives of malaria patients, with no history of fever in the past 48 h and with blood film negative for malaria parasites.

### Malaria Volunteer Infection Studies

Malaria volunteer infection studies (VIS) were conducted in Brisbane, Australia, as previously described [11]. In brief, healthy malaria-naïve volunteers were inoculated with ∼2800 viable *P. falciparum* 3D7-infected RBCs, or ∼1,800 *P. vivax*-infected RBCs. Peripheral blood parasitemia was measured by qPCR daily from day 4 until administration of antimalarial drugs, which occurred on day 7, 8 or 10, depending on the study, when parasitemia for most participants had exceeded 5,000 parasites/ml. Venous blood was collected in lithium heparin and citrate tubes for pathophysiological biomarkers prior to inoculation and prior to treatment in all studies, and at other timepoints in some of the studies (**Supplementary Table 1**). Participants with *P. falciparum* and with samples available were enrolled in studies NCT02783820 [32], NCT02389348 [33], NCT02281344 [34], and NCT02573857 [35]. Participants with *P. vivax* were enrolled in studies NCT02573857 [36] and ACTRN12616000174482 [37].

### ELISAs

For the Malaysian studies and for the malaria VIS, venous blood collected in lithium heparin and citrate tubes was centrifuged (including a second high-spin speed for the citrate tube) within 30 min of collection and plasma stored at −70° C. Plasma concentrations of OPG were measured using a duoset ELISA from RnD. Free soluble RANKL and TRAIL were measured on lithium heparin plasma by ELISA (Biomedica Diagnostics). Angiopoietin-2, P-selectin, and adhesion molecules intercellular adhesion molecule (ICAM)-1 and E-selectin were measured on lithium heparin plasma using quantikine ELISA kits from RnD. Plasma von Willibrand factor (vWF) was measured on the citrated platelet-free plasma by ELISA (Bethyl Laboratories and Biomedica Diagnostics, respectively). For logistical reasons, not all of the laboratory assays were performed on all patients or participants.

### B cell stimulation, and *ex vivo* OPG production

To assess if parasite stimulation could directly induce B cell OPG production, PBMCs were isolated from buffy coats obtained from healthy blood donors (n=10) from Australian Red Cross Lifeblood. PBMCs were isolated and fractioned by Ficoll® Paque Plus (Cytiva) and frozen in 10% DMSO with 90% FBS. PBMCs were thawed in 10% FBS/RPMI, and 1 million PBMCs stimulated with 3 million parasitized red blood cells (pRBCs) or unparasitised RBCs (uRBCs) for 3 days. Total B cells were characterised as CD19+ and B cell subsets identified by IgD, CD21 and CD27. In brief, 1 million PBMCs were surface stained with NIR Live/Dead (Thermofisher), CD19-BUV395 (SJ25C1, 1/50), CD3-FITC (HIT3a, 1/25), CD21-PE-Dazzle (Bu32, 1/50), CD27-BV421 (M-T271, 1/50), IgD-PE-Cy7 (IA6-2, 1/50), CD14-BV650 (M5E2, 1/50) and CD56-BV510 (HCD56, 1/25) for 30 minutes at room temperature followed by 2% FBS/PBS wash. Cells were fixed and permeabilized using 1X Perm/Wash^TM^ (BD Biosciences, USA) and incubated with anti-human polyclonal OPG-biotin (1/15 Leinco technologies, USA) at 30 minutes at room temperature, followed by the addition of Streptavidin-PE (0.125 mg per million cells) to quantify intracellular OPG. All cells were acquired on a Fortessa V flow cytometer (BD Immunocytometry Systems, San Jose, CA) and the data was analysed using FlowJo software version 10 (Treestar, San Carlos CA). Surface antibodies were sourced from BD Biosciences (USA) and Biolegend (USA).

To assess if B cell OPG production was increased in clinical malaria, PBMCs from Malaysian patients with malaria and from healthy controls from the same study site were thawed in RPMI containing 10% FBS and rested overnight at 37°C, 5% CO2. Cells were then analysed by flow cytometry as for parasite stimulated cells.

### Ethics Statement

All studies were conducted in accordance with the Declaration of Helsinki and the International Committee of Harmonisation of Good Clinical Practice guidelines. All participants provided informed written consent. For the use of samples from the malaria volunteer infection studies and for the isolation of PBMCs from buffy coats obtained from Australian Red Cross Lifeblood, ethics approval was obtained from the Human Research Ethics Committee of the QIMR Berghofer Medical Research Institute (P1479 and P3444 respectively). For the clinical studies in Malaysia, ethics approval was obtained from the Malaysian Ministry of Health (NMRR10_754_6684) and the Menzies School of Health Research, Darwin, Australia (HREC 2010_1431). Informed written consent was provided by all participating adults and parents/guardians of those under 18 years of age.

### Role of Funders

The funders had no role in the study design, data collection, data analysis, interpretation or writing of this report.

### Statistical analysis

Statistical analyses were performed using Stata software (version 15) and GraphPad Prism 8.4.3. For continuous variables, comparisons between groups were analysed using student’s T-test or Mann-Whitney tests, depending on distribution. Categorical variables were compared using χ^2^ test. Associations between continuous variables were assessed using Spearman’s or Pearson’s correlation coefficients, depending on distribution. Partial correlations were used to adjust for parasitaemia and age, with non-normally distributed variables log-transformed to normality. A backwards stepwise regression model was used to evaluate the association between TRAIL and acute kidney injury (defined by KDIGO criteria), with variables log transformed and removed if p>0.05. Wilcoxon signed rank test was used to compare baseline and follow-up measurements. Measurements below the limit of detection were assigned a value of half the lower limit of detection.

## Data Availability statement

The datasets generated during and/or analysed during the current study are available from the corresponding author on reasonable request.

## Declaration of Interests

None of the authors have conflicts of interest to declare.

## Acknowledgements

We thank all volunteers enrolled in the malaria volunteer infection studies in Brisbane, Australia, and the patients enrolled in the clinical studies in Malaysia; research and clinical staff at the participating hospitals in Malaysia; and clinical staff at QPharm Clinical Trial Site, Brisbane, Australia.

This study was supported by the National Health and Medical Research Council (Program Grant 1037304, Project Grants 1045156 and 1156809; Investigator Grants 2016792 to BEB, 2016396 to JSM, 2017436 to MJG), the US National Institute of Health (R01 AI116472-03), and the Malaysian Ministry of Health (Grant BP00500420). The malaria volunteer infection studies are supported by Medicines for Malaria Venture.

## Author Contributions

N.M.A., M.J.B and B.E.B. conceived and designed the study. B.E.B, M.J.G, and T.W. conducted the clinical studies in Malaysia. J.W., B.E.B., and J.S.M, conducted the malaria volunteer infection studies. K.P. conducted the ELISAs. A.S.N, D.A., J.L. and F.A. conducted the B cell assays. A.S.N, J.W., J.L., M.J.B and B.E.B. analysed the data. A.S.N, J.W., J.L, M.J.B and B.E.B. wrote the paper, with input from all other authors. All authors approved the final draft of the manuscript.

## Notes

### Competing Interest Statement

The authors have declared no competing interest.

### Funding Statement

This study was funded by the National Health and Medical Research Council (Program Grant 1037304, Project Grants 1045156 and 1156809; Investigator Grants 2016792 to BEB, 2016396 to JSM, 2017436 to MJG), the US National Institute of Health (R01 AI116472-03), and the Malaysian Ministry of Health (Grant BP00500420). The malaria volunteer infection studies were supported by Medicines for Malaria Venture.

