## Supplementary Data for "Osteoprotegerin (OPG) and its ligands RANKL and TRAIL in falciparum, vivax and knowlesi malaria: correlations with disease severity, and B cell production of OPG"

**Supplementary Table 1. Logistic regression model for predicting acute kidney injury in patients with knowlesi malaria.**

| | Odds Ratio | 95% Confidence Interval | P value |
|---|---|---|---|
| **Predictors of AKI:** | | | |
| Log osteoprotegerin | 2.74 | 1.50 – 5.00 | 0.001 |
| Log TRAIL | 0.28 | 0.11 – 0.73 | 0.009 |
| Age | 1.07 | 1.04 – 1.10 | <0.001 |
| **Alternative model, including Ang2:** | | | |
| Log Ang2 | 4.39 | 2.01 – 9.55 | <0.001 |
| Log osteoprotegerin | 1.97 | 1.02 – 3.80 | 0.044 |
| Age | 1.07 | 1.04 – 1.10 | <0.001 |

AKI: acute kidney injury as defined by KDIGO; Ang2: angiopoietin-2; TRAIL: tumour necrosis factor-related apoptosis inducing ligand. Backward stepwise regression was used, with variables removed if P value was >0.05. Variables included in both models included: age, angiopoietin-2, osteoprotegerin, TRAIL and parasite count.

**Supplementary Table 2. Correlations with OPG/TRAIL ratio in patients with falciparum, vivax and knowlesi malaria.**

| | *P. falciparum* (n=169) | | | | *P. vivax* (n=61) | | | | *P. knowlesi* (n=197) | | | |
|---|---|---|---|---|---|---|---|---|---|---|---|---|
| | Univariate analysis | | Controlling for parasitaemia and age | | Univariate analysis | | Controlling for parasitaemia and age | | Univariate analysis | | Controlling for parasitaemia and age | |
| | Correlation coefficient | P value | Correlation coefficient | P value | Correlation coefficient | P value | Correlation coefficient | P value | Correlation coefficient | P value | Correlation coefficient | P value |
| Parasite count | 0.34 | <0.0001 | 0.34 | <0.0001[a] | 0.45 | 0.0001 | | | 0.49 | <0.0001 | 0.43 | <0.0001[a] |
| Age | 0.27 | 0.0003 | 0.22 | 0.0036[b] | 0.17 | 0.21 | | | -0.41 | <0.0001 | 0.30 | <0.0001[b] |
| HRP2[c] | 0.05 | 0.50 | | | | | | | | | | |
| Creatinine | 0.25 | 0.0009 | 0.13 | 0.11 | 0.12 | 0.35 | | | 0.49 | <0.0001 | 0.43 | <0.0001 |
| Lactate | 0.39 | <0.0001 | 0.28 | 0.0007 | 0.24 | 0.077 | | | 0.37 | <0.0001 | 0.32 | <0.0001 |
| IL-6 | 0.23 | 0.0066 | 0.37 | <0.0001 | 0.52 | <0.0001 | 0.41 | 0.0013 | 0.57 | <0.0001 | 0.32 | <0.0001 |
| Ang-2 | 0.32 | <0.0001 | 0.24 | 0.0015 | 0.45 | 0.0004 | 0.28 | 0.032 | 0.60 | <0.0001 | 0.49 | <0.0001 |
| vWF[d] | 0.34 | 0.012 | 0.07 | 0.64 | 0.42 | 0.010 | 0.21 | 0.24 | 0.27 | 0.015 | 0.23 | 0.039 |
| P-selectin | 0.08 | 0.31 | | | 0.12 | 0.38 | | | 0.22 | 0.0016 | 0.17 | 0.016 |
| ICAM-1 | 0.19 | 0.011 | 0.27 | 0.0003 | 0.09 | 0.48 | | | 0.29 | <0.0001 | 0.23 | 0.0016 |
| E-selectin | 0.41 | <0.0001 | 0.37 | <0.0001 | 0.38 | 0.0030 | 0.32 | 0.014 | 0.29 | <0.0001 | 0.23 | 0.0011 |
| Hb nadir | 0.13 | 0.082 | | | -0.02 | 0.85 | | | -0.35 | <0.0001 | -0.22 | 0.0023 |
| Plt nadir | -0.34 | <0.0001 | -0.34 | <0.0001 | -0.19 | 0.14 | | | -0.50 | <0.0001 | -0.29 | 0.0001 |
| RHPAT[e] | 0.03 | 0.77 | | | -0.13 | 0.45 | | | -0.41 | <0.0001 | -0.26 | 0.010 |

[a]controlling for age; [b]controlling for parasitaemia; [c]measured in 158 patients; [d]measured in 54 patients with *P. falciparum*, 37 with *P. vivax* and 85 with *P. knowlesi*; [e]measured by peripheral arterial tonometry in 88 patients with *P. falciparum* and 38 patients with *P. vivax*.. HRP2: histidine rich protein 2; IL-6: interleukin 6; Ang-2: angiopoietin-2; vWF: von Willebrand Factor; OPG: osteoprotegerin; RANKL: receptor activator of nuclear factor kappa B lamda; ICAM-1: intracerebral adhesion molecule 1; Hb: haemoglobin; Plt: platelet.

**Supplementary Table 3: Baseline characteristics of IBSM participants by study**

| Study | NCT02783820 | NCT02389348 | NCT01055002 | ACTRN12616000174482 | NCT02573857 |
|---|---|---|---|---|---|
| Cohort | 5 | 1 | 2, 3 | 1 | 2 |
| Year | 2016 | 2015 | 2010 | 2016 | 2016 |
| Number of participants | 8 | 7 | 13 | 8 | 8 |
| Inoculum species | *P. falciparum* | *P. falciparum* | *P. falciparum* | *P. vivax* | *P. vivax* |
| Inoculum size (number of viable parasites) | 2800 | 1800 | 1800 | 564 | 564 |
| Day of treatment (days post inoculum) | 8 | 7 | 8 | 8 | 10 |
| Day of End of Study sample collection (days post inoculum) | 14 | NA | 28 | 28 | 28 |
| Age (years; median, range) | 29 (20-48) | 24 (19-55) | 26 (19-43) | 19.5 (18-30) | 23 (21-31) |
| Sex (male) | 8/8 | 4/7 | 13/13 | 5/8 | 8/8 |
| Parasite count on day of treatment (parasites/mL, median, range) | 11,535 (1,508-95,851) | 1,751 (289-7,678) | 803 (219-13,847) | 5,737 (1,288-9,209) | 80,149 (30,331-175,522) |
| **Days of sample collection** | | | | | |
| OPG | 0, 5, 6, 7, 8, 14 | 0, 7 | 0, 3, 4, 5, 6, 7, 8, 9, 10, 28 | 0, 5, 6, 7, 8, 15, 28 | 0, 5, 6, 7, 8, 9, 10, 18 |
| RANKL | 0, 7, 8, 14 | 0, 7 | 0, 7, 8, 9, 10, 28 | 0, 7, 8, 15, 28 | 0, 7, 8, 9, 10, 18, 28 |
| TRAIL | 0, 5, 6, 7, 8, 14 | 0, 7 | 0, 3, 4, 5, 6, 7, 8, 9, 10, 28 | 0, 5, 6, 7, 8, 15, 28 | 0, 5, 6, 7, 8, 9, 10, 18 |
| vWF | 0, 5, 6, 7, 8 | 0, 7 | NA | 0, 5, 6, 7, 8 | 0, 5, 6, 7, 8, 9, 10 |
| Ang2 | 0, 5, 6, 7, 8 | 0, 7 | NA | 0, 5, 6, 7, 8 | 0, 5, 6, 7, 8, 9, 10 |
| Hematology | 0, 8 | 0, 7 | NA | 0, 8 | 0, 10 |

NA = not applicable; OPG = osteoprotegerin; RANKL = receptor activator of NFkB ligand; TRAIL = TNF-related apoptosis inducing ligand; vWF = von Willebrand factor; Ang2: angiopoietin 2

**Supplementary Table 4: Correlations with RANKL and other selected variables in participants enrolled in *P. falciparum* and *P. vivax* malaria volunteer infection studies, on day of treatment.**

| | *P. falciparum* (n=28) | | | | *P. vivax* (n=16) | | | |
|---|---|---|---|---|---|---|---|---|
| | Univariate analysis | | Controlling for parasitaemia | | Univariate analysis | | Controlling for parasitaemia | |
| | Correlation coefficient | P value | Correlation coefficient | P value | Correlation coefficient | P value | Correlation coefficient | P value |
| OPG | -0.66 | 0.0002 | -0.63 | 0.0004 | -0.92 | <0.0001 | -0.76 | 0.0011 |
| TRAIL | -0.50 | 0.031 | -0.57 | 0.013[a] | -0.63 | 0.012 | -0.01 | 0.96 |
| Parasitemia | 0.07 | 0.71 | | | -0.84 | 0.0001[a] | | |
| vWF | 0.01 | 0.98 | | | -0.64 | 0.0088 | -0.42 | 0.12 |
| Angiopoietin-2 | 0.17 | 0.53 | | | -0.48 | 0.063 | | |
| Hemoglobin | 0.11 | 0.69 | | | -0.17 | 0.53 | | |
| Platelet count | 0.23 | 0.40 | | | 0.45 | 0.082 | | |

OPG = osteoprotegerin; TRAIL = TNF-related apoptosis inducing ligand; vWF = von Willebrand factor; RANKL = receptor activator of NFkB ligand. [a]did not remain significant after controlling for OPG.

**Supplementary Table 5: Correlations with TRAIL and other selected variables in participants enrolled in *P. falciparum* and *P. vivax* malaria volunteer infection studies, on day of treatment.**

| | *P. falciparum* (n=28) | | | | *P. vivax* (n=16) | | | |
|---|---|---|---|---|---|---|---|---|
| | Univariate analysis | | Controlling for parasitaemia | | Univariate analysis | | Controlling for parasitaemia | |
| | Correlation coefficient | P value | Correlation coefficient | P value | Correlation coefficient | P value | Correlation coefficient | P value |
| OPG | 0.22 | 0.37 | | | 0.64 | 0.0089 | 0.05 | 0.87 |
| Parasitemia | 0.03 | 0.91 | | | 0.68 | 0.0048 | | |
| vWF | -0.04 | 0.89 | | | 0.64 | 0.0095 | 0.25 | 0.37 |
| Angiopoietin-2 | -0.27 | 0.33 | | | 0.46 | 0.078 | | |
| Hemoglobin | 0.49 | 0.065 | | | 0.36 | 0.17 | | |
| Platelet count | -0.8 | 0.026 | -0.60 | 0.023[a] | -0.65 | 0.0082 | -0.40 | 0.14 |

OPG = osteoprotegerin; TRAIL = TNF-related apoptosis inducing ligand; vWF = von Willebrand factor; RANKL = receptor activator of NFkB ligand. [a]remained significant after controlling for OPG.

**Supplementary Table 6: Patient demographics for cell samples used to assess OPG during malaria**

| | *P. falciparum* (n=6) | *P. vivax* (n=8) | *P. knowlesi* (n=7) | Healthy controls (n=5) |
|---|---|---|---|---|
| Age, years (median, range) | 38 (27-54) | 39 (16-54) | 48 (25-55) | 30 (22-36) |
| Male sex, n (%) | 3 (50) | 6 (75) | 6 (86) | 4 (80) |
| Parasitemia (parasites/µL, median, range) | 30,086 (1508-273,908) | 14048 (1464-84403) | 292007 (13,680-584,015) | NA |

**Supplementary Figure 1. Longitudinal measurements of RANKL in Malaysian patients with *P. knowlesi* (A), *P. falciparum* (B) and *P.vivax* (C) malaria. D: RANKL concentrations at day 28 compared to controls.** In panel D, error bars represent median and inter-quartile range.

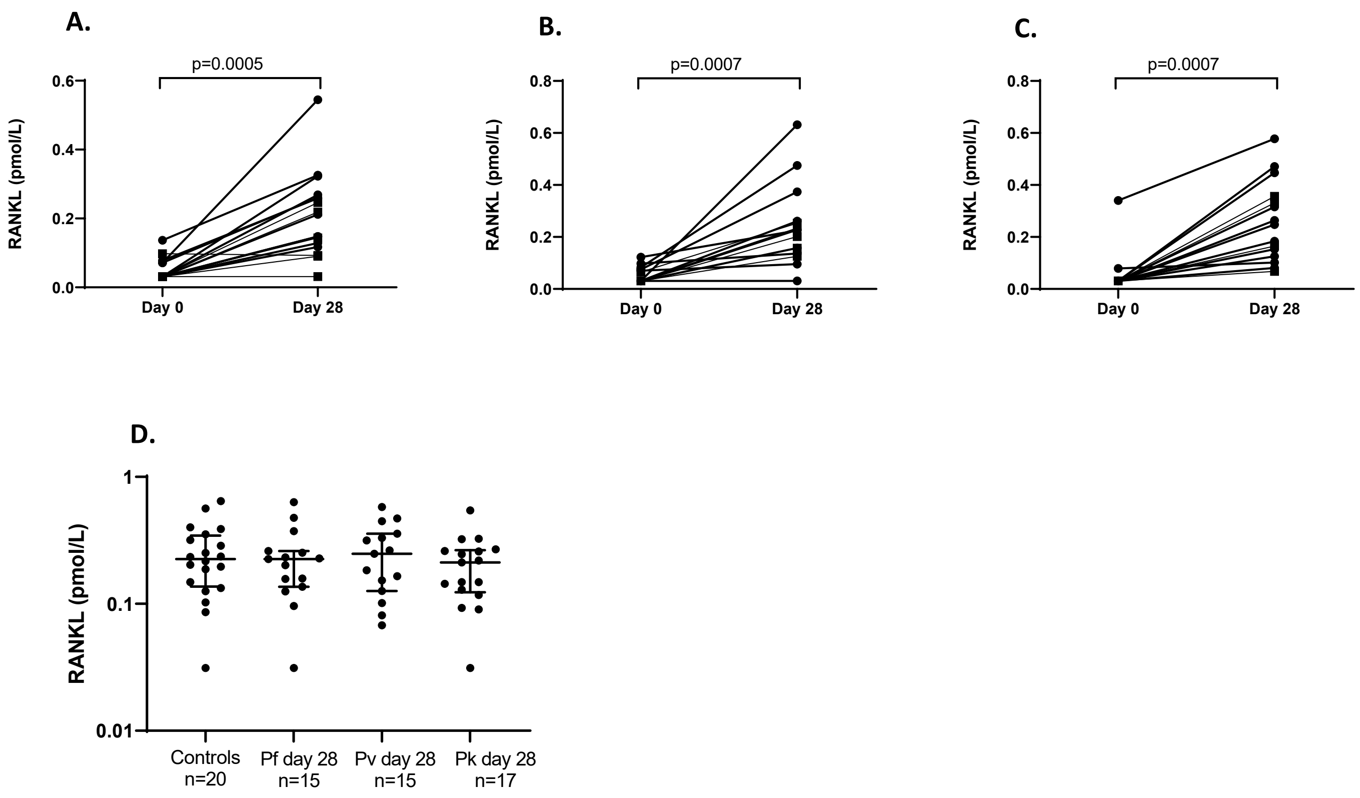

**Supplementary Figure 2. Other cell subsets do not express OPG after in vitro *Plasmodium* stimulation in malaria naive donors. A)** Gating strategy example of B cell subsets, T cells, NK cells and monocytes by flow cytometry. **B)** Flow cytometry example of OPG expression from T cells, NK cells and Monocytes at enrolment after uRBC (grey) and pRBC (green) in vitro stimulation.

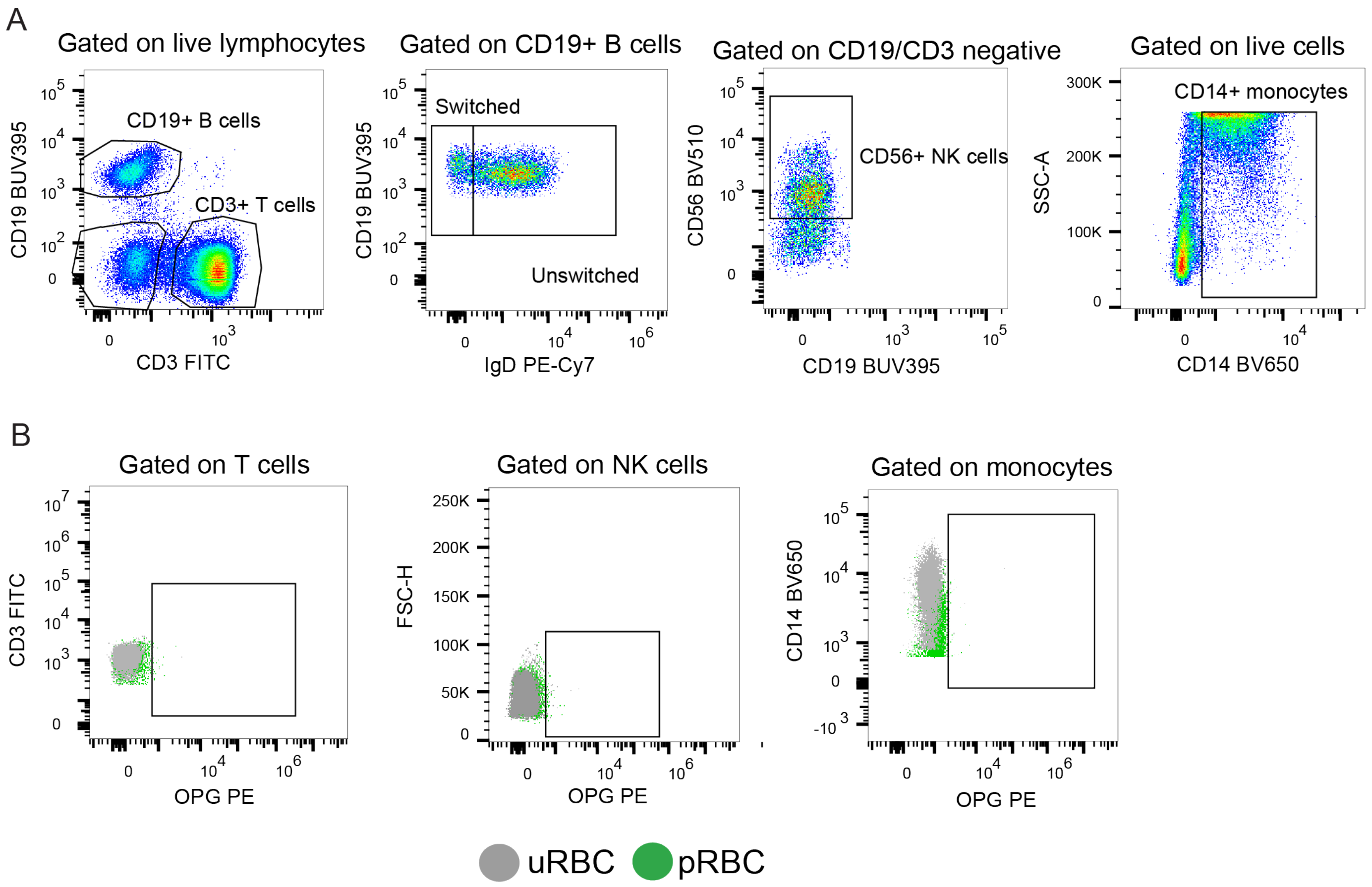

**Supplementary Figure 3. Production of OPG from other cell subsets during clinical malaria. A)** Gating strategy example of B cell subsets, T cells, NK cells and monocytes by flow cytometry. **B)** Flow cytometry example of OPG expression from T cells, NK cells and Monocytes at enrolment (day 0), example plot is from a *P. knowlesi* patient.

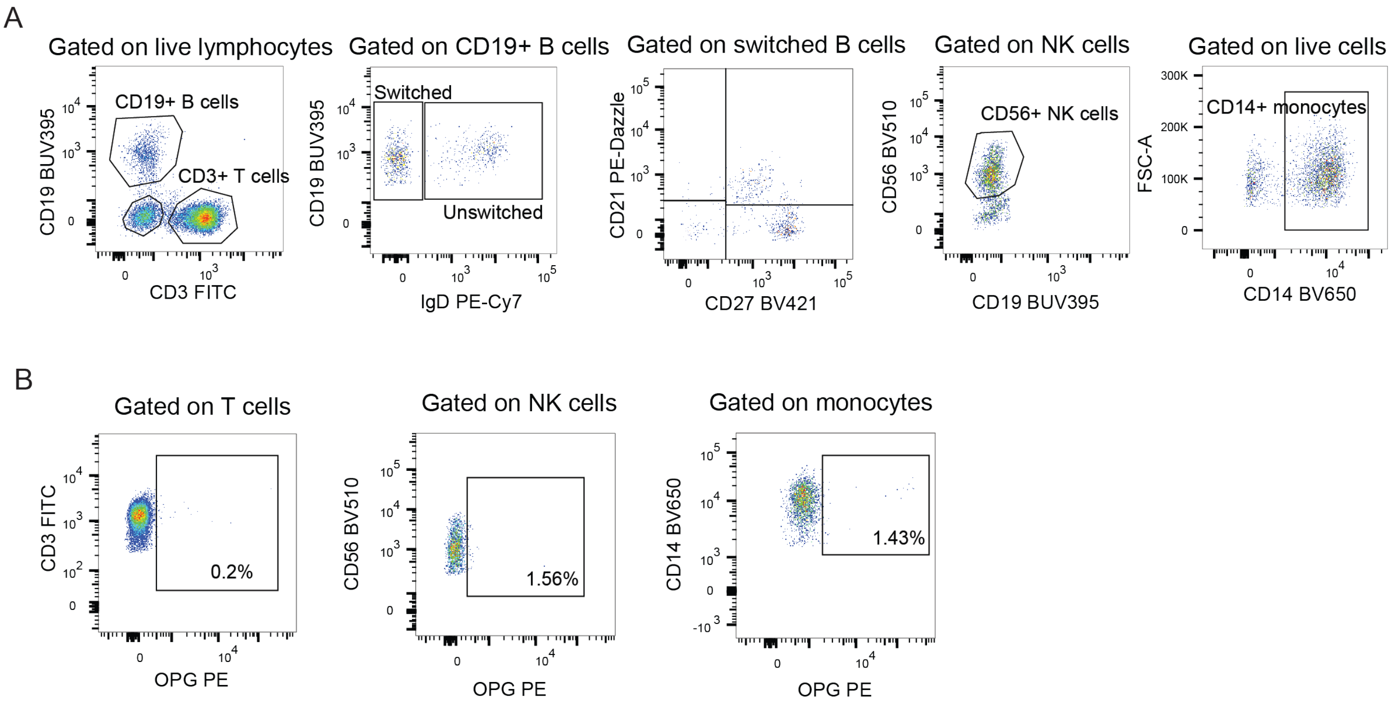
